# Cardiovascular risk in a rural psychiatric inpatient population: Retrospective case cohort study

**DOI:** 10.1101/2022.09.11.22279816

**Authors:** Alan Woodall, Amy Prosser, Millie Griffiths, Ben Shooter, Joy Garfitt, Lauren Walker, Iain Buchan

**Affiliations:** Mental Health Directorate, Bronllys Hospital, Powys Teaching Health Board, Brecon LD3 OLW and Honorary Clinical Research Fellow, Department of Primary Care and Mental Health, Institute of Population Health, University of Liverpool, L69 3GF; Mental Health Directorate, Bronllys Hospital, Powys Teaching Health Board, Brecon LD3 OLW, L69 3GF; Mental Health Directorate, Bronllys Hospital, Powys Teaching Health Board, Brecon LD3 OLW; Institute of Systems, Molecular and Integrative Biology, University of Liverpool, L69 7DE; Institute of Population Health, University of Liverpool, L69 3GF

## Abstract

**Aims and Method:** To evaluate cardiovascular risk in a rural inpatient psychiatric unit over a one-year period. Care records were analysed for risk factor recording, and cardiovascular risks estimated using the QRISK3 calculator, which estimates 10-year risk of myocardial infarction or stroke.

**Results:** Of eligible patients, risk factor recording as part of routine care was completed in 86% of possible QRISK3 inputs, enabling QIRSK3 estimation in all eligible patients. QRISK3 for this group was raised (Relative risk: 3.8, 95%CI: 2.5 – 5.0). High risk of cardiovascular disease (QRISK3 score >10%) was detected in 28% of patients.

**Clinical Implications:** This service evaluation demonstrated significant unmet need for cardiovascular risk reduction that could be identified as part of routine care. An opportunity exists to integrate mental and physical healthcare by routinely assessing cardiovascular risk in psychiatric inpatients. Resources and training are needed to produce this risk information and act on it.

## INTRODUCTION

Patients with serious mental illness (SMI) such as schizophrenia and bipolar affective disorder die 15-20 years earlier than the general population (1-3); the leading cause is cardiovascular mortality (4, 5). This is due to genetic, behavioural, socioeconomic, and service organisational factors (5-7), in addition to increased risk from atypical antipsychotics prescribed to most patients (8, 9). Modifiable factors account for two-thirds of the attributable risk fraction for this population (10). SMI patients are often admitted as inpatients for prolonged periods, providing unique opportunities to ameliorate these cardiovascular risks (11, 12). Improving the physical health of people living with SMI is a priority for UK Governments, with toolkits to support interventions (13).

Quantifiable estimation of cardiovascular risk is facilitated by online calculators that produce a risk score from commonly recorded variables. QRISK3 is recommended by the NHS as a validated cardiovascular risk tool (14) and NICE recommends consideration of lipid-lowering therapy for anyone with a ‘high’ risk score >10% (15). QRISK3 calculates the 10-year risk of experiencing a myocardial infarction or stroke for the average individual with the given input characteristics, using up to twenty-two risk factors. QRISK3 estimation allows shared decision-making to address modifiable risk factors, including behaviour change (e.g., smoking cessation) and pharmacological intervention to reduce physiological risks, such as commencing lipid-lowering therapy, or changing antipsychotic to reduce risk.

In this study, we evaluate QRISK3 input data completeness for patients admitted to the inpatient unit to have their cardiovascular risk assessed, informing interventions as needed.

## METHODS

### Setting

Patients admitted to the psychiatric general adult service at Bronllys Hospital, Powys from May 1^st^ 2021 to 30^th^ April 2022. The inpatient unit has eighteen beds for adults aged 18-65 years, serving a highly rural and sparsely populated part of Wales.

### Record analysis and selection criteria

Records were analysed for demographic information (age, sex, ethnicity, postcode) and whether admission was voluntary or via detention under the Mental Health Act (1983) (MHA). QRISK3 data were extracted from clinical records, including mental health diagnosis on discharge, and whether any family or past medical history of physical health disease was noted that affects QRISK3 calculation, including any pre-existing cardiovascular disease. The admission pathway requests a GP summary for the patient, which was analysed for any current investigation data, and if recorded in the prior 12 months, used for the QRISK3 calculation. Current medications, mental health diagnosis, and physical health parameters recorded during admission including smoking status, weight, height, body mass index (BMI), admission blood pressure, and blood investigations relevant to physical health outcomes (cholesterol/High Density Lipoprotein (HDL) ratio, HBA1c) were extracted. We did not attempt to calculate standard deviations for at least two blood pressure readings (an option in the QRISK3 calculator) as this did not represent a practical option for most care contexts – the data are not commonly available. Thus, an estimate of the number of twenty-one possible QRISK3 risk factors were derived. Where a parameter was missing or not recorded that would affect QRISK3 calculation, (e.g., a record of family history of early cardiovascular disease, such as a myocardial infarction or angina before the age of 60), this was omitted from QRISK3 calculation, or the assumed lowest risk profile selected where not recorded (e.g., ‘never smoked’). Where smoking status was only recorded as ‘YES’ or ‘NO’, rather than categorised as ‘never-, ex-, light-, moderate- or heavy-smoker’ as QRISK3 requires as a mandatory factor for calculation, those recorded ‘YES’ were assigned a ‘moderate smoking (10-19 cigarettes a day)’ status, and if ‘NO’ assigned a ‘never smoked’ status. Studies (16) of psychiatric inpatients suggest that up to 70 per cent smoke, and around 50 per cent are heavy smokers (>20 cigarettes per day); this method to address missing or incomplete smoking risk factor record was thus felt a reasonable assumption for calculation purposes, ensuring the QRISK3 calculations would be the lowest possible risk estimation in non-smoking cases, and a pragmatic estimate of cigarette consumption in smoking patients if not recorded in more detail. The number of missing QRISK3 variables for each patient were recorded, and all results also analysed by sex and MHA status.

Patients were excluded from analysis if they were outside the age range (25 - 84 years) that QRISK3 provides estimates for. If a patient had an admission duration of 5 days or less, they were also excluded from analysis, as this was felt insufficient time to have appropriate work-up and intervention, including blood tests, record retrieval and staff time to discuss cardiovascular risk with the patient pragmatically. Patients with known pre-existing cardiovascular disease were also excluded, as QRISK3 is a primary prevention calculator and is not validated for secondary prevention or risk estimate. We also recorded if the patient had an estimate of HBA1c to assess potential diabetic risk, and to confirm recorded diabetic status for the calculation. Where individuals had multiple admissions during the study period, we used the highest QRISK3 score from one admission for summary analysis.

### Statistical methods

Statistical analysis was performed using SPSS (version 28). For two-by-two binary variable contrasts a two-sided Fisher exact P is given. For continuous variables with approximately symmetrical distributions, means are compared with a two-tailed Student’s t-test. A significant difference was assumed if P<0.05. Results are presented as main effect with a 95% confidence interval where possible.

### Ethical approval

As a retrospective service improvement evaluation of routinely collected patient records, ethical and governance approval was granted by Powys Teaching Health Board Research and Development (R&D) directorate (approvals recommendation: PTHB-21) Consent was not sought for this audit of routinely obtained data from patient records for service improvement purposes.

## RESULTS

### Admissions analysis, assessment of QRISK3 variable recording and individual QRISK3 score calculation

Figure 1 shows the pathway for inclusion for QRISK3 assessment. There were 168 separate admission events in the study period; four events were unobtainable, and four events were readmission to the unit following an interim transfer to a psychiatric intensive care unit, so counted as a single admission. Of the 160 admissions reviewed, 62 were admissions that <= 5 days, 18 admissions were below age threshold for QRISK3 (<25 years), and 5 admissions were for patients with known cardiovascular disease; these 85 events were excluded from QRISK3 analysis. Seventy-five admission events were thus eligible for QRISK3 assessment, and fifty-seven individual patients had a QRISK3 estimation undertaken (sixteen patients had two eligible admissions, and two patients had three eligible admissions during the study period).

**Figure 1:**
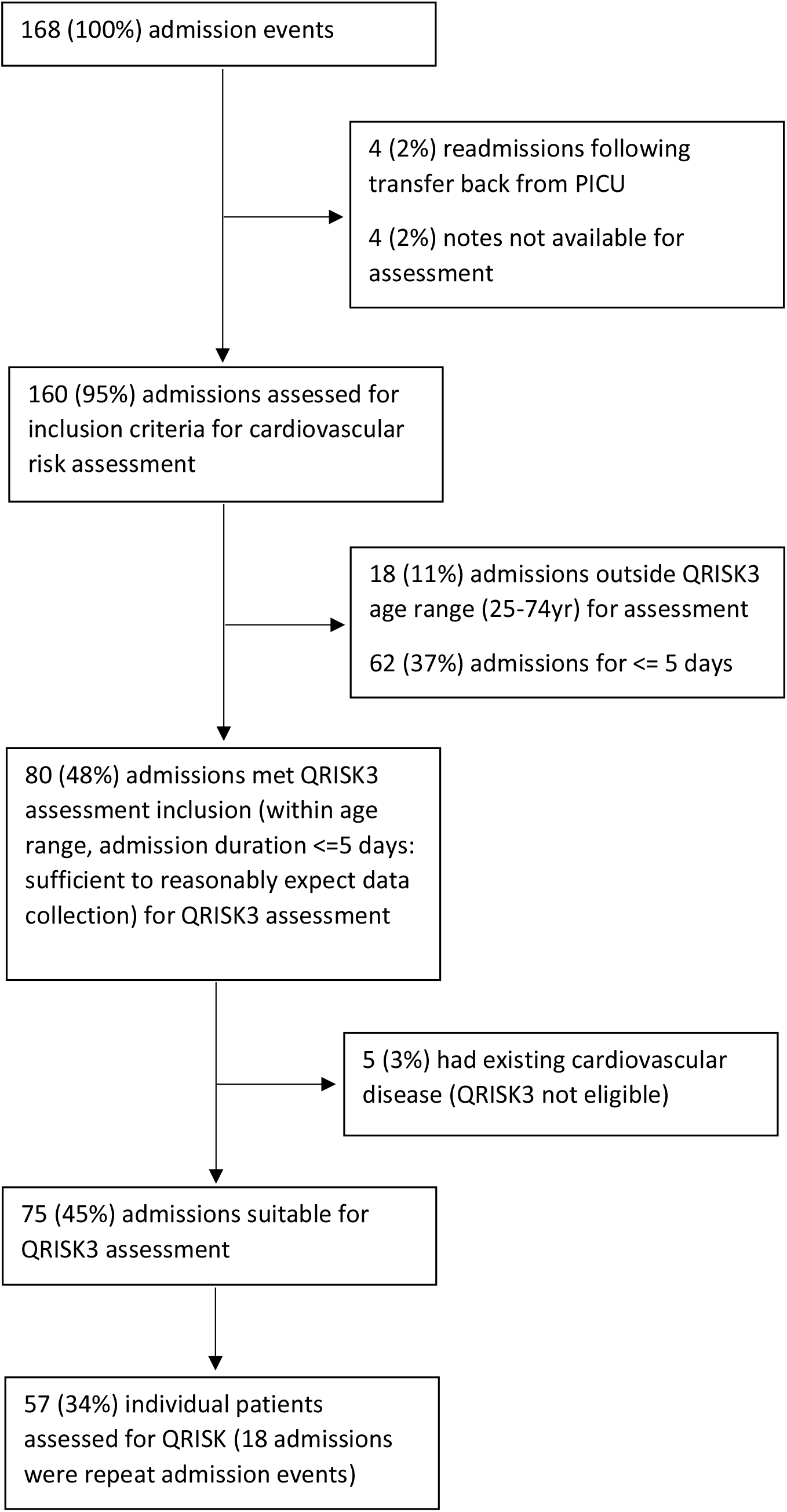
Flowchart of admissions assessed for QRISK3 estimation.

### Recording of risk variables to enable QRISK3 cardiovascular assessment

Table 1 shows the risk factor capture for the 75 QRISK3 assessments undertaken for each admission event. The mean admission event duration was 49 days; mean age was 44.6 years; males comprised 48% of admissions; 93% of admissions were ‘White-British’ ethnicity and 71% of admissions involved detention under the Mental Health Act.

**Table 1:**
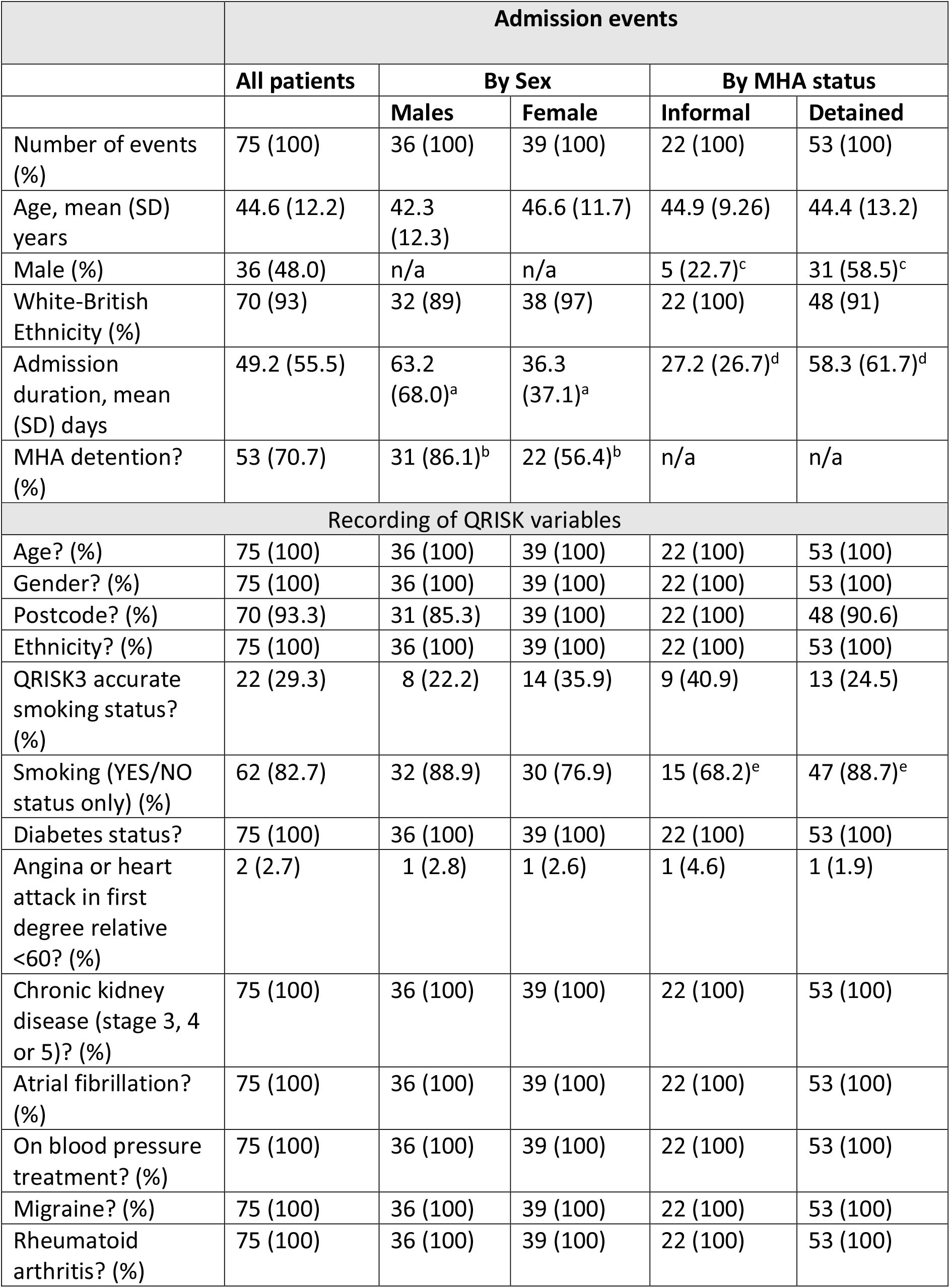
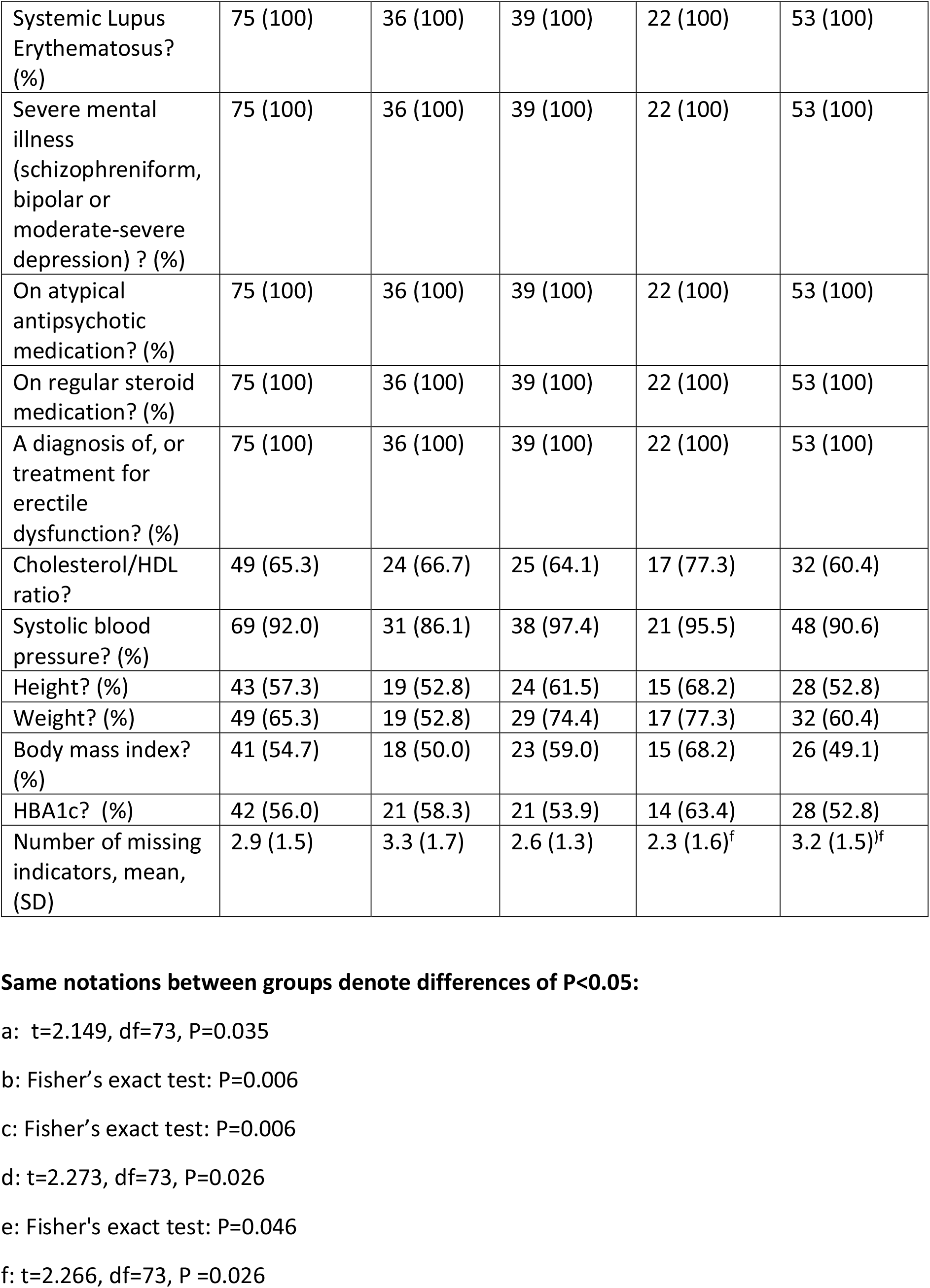
Recording accuracy of QRISK3 variables to allow QRISK 3 assessment.

Recording of risk factors to enable a QRISK3 assessment was highly variable and depended on the source; many variables are routine data collected during admission and had 100% recording (age, sex, ethnicity) and others are collated from GP records obtained electronically for each GP practice (past medical history, current medications), again allowing in most cases near 100% capture of variables. Postcode was available for nearly all patients (93%), but a number were of no fixed abode; for these, the postcode for the most deprived lower super-output area in the 2019 Welsh Index of Multiple Deprivation Index (LL18 1AB) was used as proxy for the calculation, as homeless patients are highly deprived.

However, a small number of factors were not readily available or rarely recorded (e.g., family history of cardiovascular disease <60 years in a first degree relative) as this is not often recorded in GP summaries and is not a question asked on admission proforma; only 3% of admissions had a record of early cardiovascular disease in family members. Recording of physical examination findings was also variable: body mass index (BMI) was recorded in just over half (55%) of patients. Blood pressure was usually obtained during admission (92% recorded). Investigations to assess lipid status were completed (or available online) for two-thirds of admissions (65%) and just over half for HBA1c (56%). Overall, the mean number of missing indicators was 2.9 (95% CI 2.6 – 3.3) out of a total possible of twenty-one separate variables that could be included in the QRISK3 calculation; thus, on average, 86% of QRISK3 variables were routinely available to undertake an estimate.

There were no significant differences between sexes for QRISK3 variable recording, despite males having longer inpatient admissions than females (difference = 26.9 (2.0 – 51.9) days, P =0.04) and being more likely to be detained under the Mental Health Act (MHA) (86% male admissions vs 56% female admissions (difference = 30% (11% – 49%), P = 0.006)).

When analysed by MHA status, there were no significant differences in QRISK3 variable recording, apart from smoking status recording was lower in voluntary admissions (68%) compared with those detained (89%) although the difference was borderline (difference =21% (0% – 42%), P =0.05). Overall, however, there were more QRISK3 variables missing during detained than voluntary admission events (difference = 0.85 (0.11 - 1.60), P = 0.03).

### Estimation of QRISK3 scores for individual patients

Table 2 shows the prevalence or mean values of QRISK3 risk factors for the fifty-seven individuals who underwent QRISK3 assessment. Mean inpatient duration was 52 days; mean age was 43 years; 51% were males; 95% white-British ethnicity and 70% were detained under the MHA. Many patients had high prevalence of modifiable risk factors: 63% were smokers, 63% had an ICD-10 coding of schizophreniform, bipolar disorder or moderate-severe depressive disorders on discharge summary, and 77% were prescribed atypical antipsychotics. Overall, the mean number of missing QRISK3 variables was 2.9 (95%CI 2.5 – 3.3) out of a maximum of twenty-one variables.

**Table 2:**
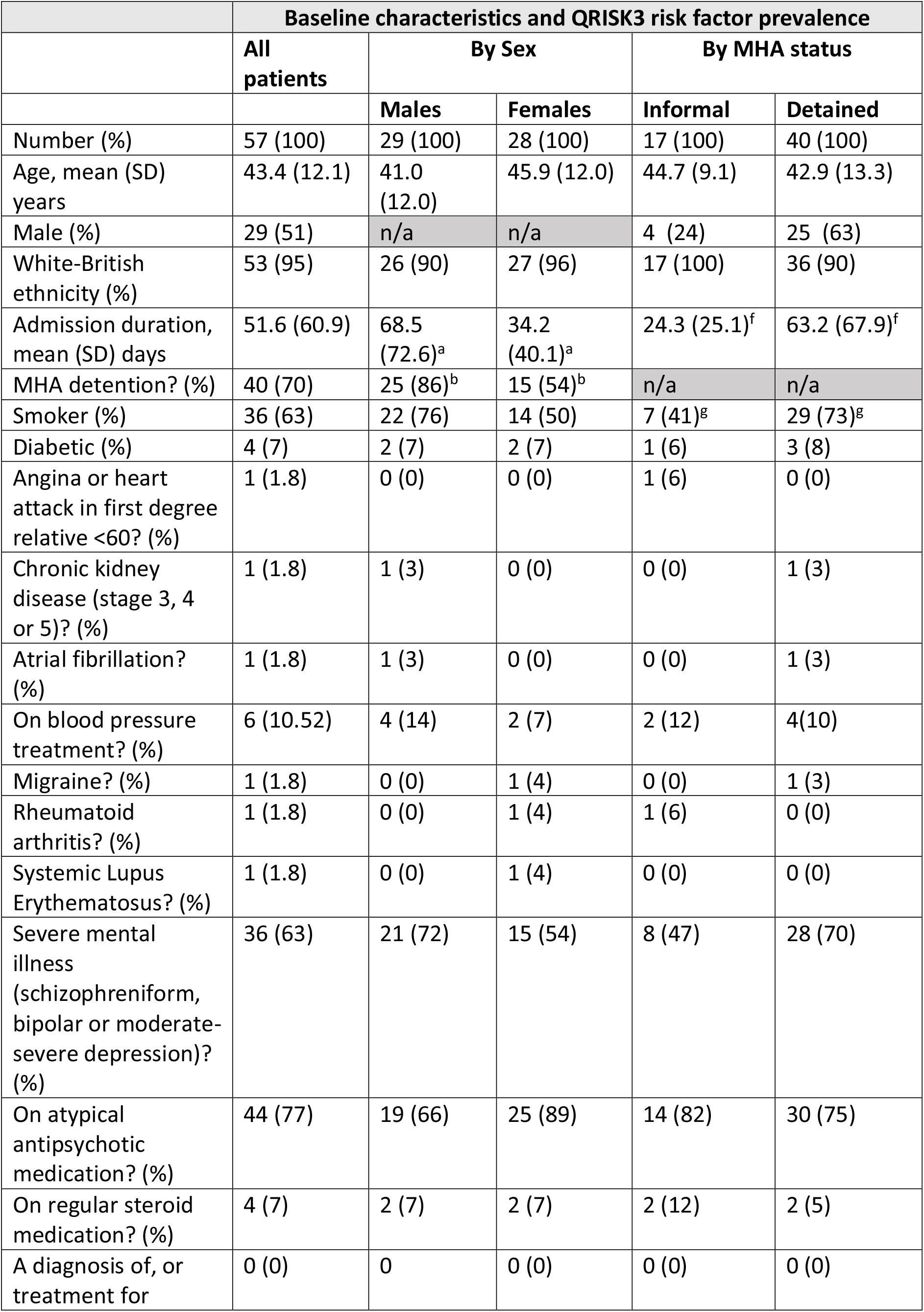
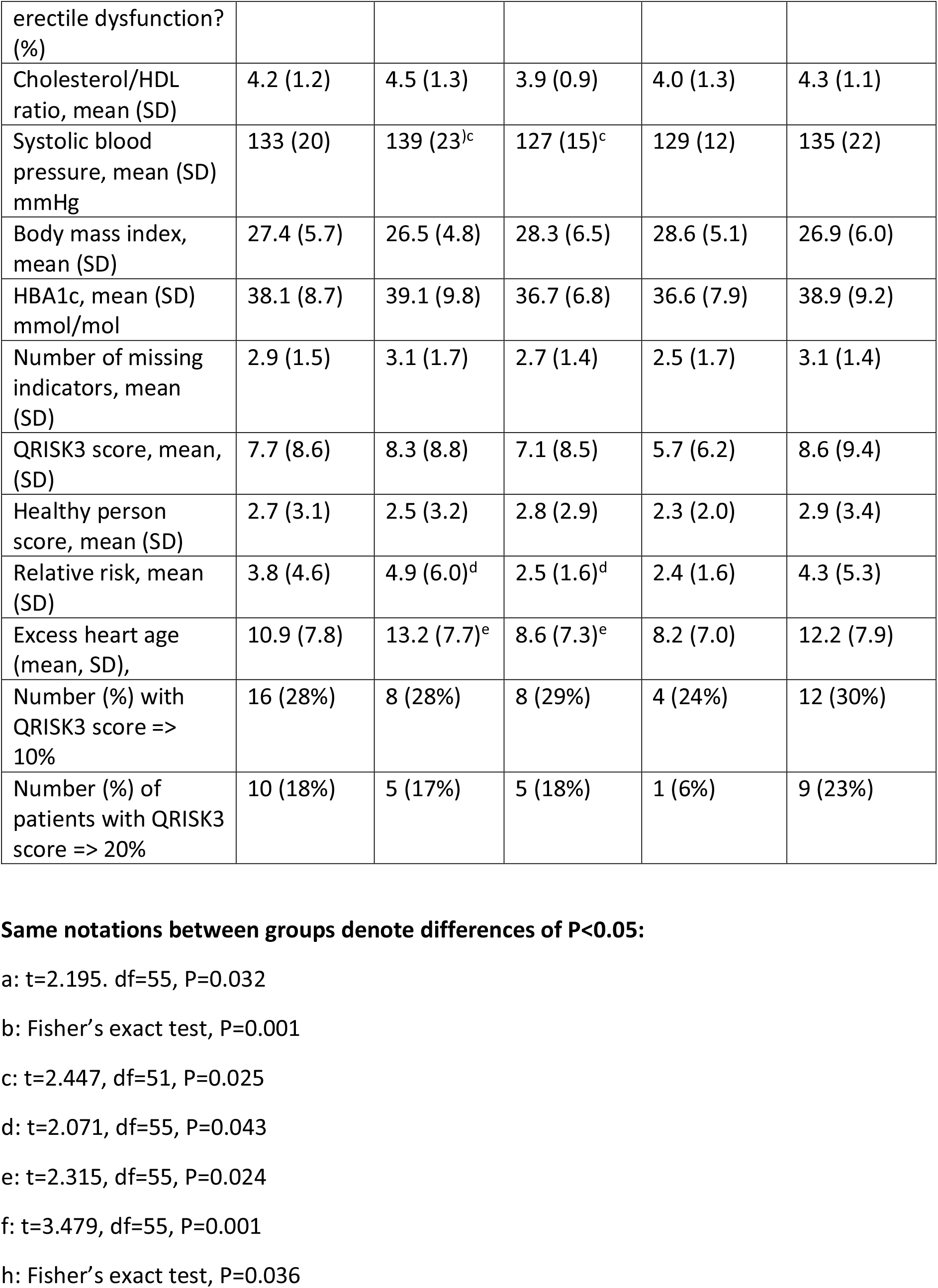
QRISK3 assessments of eligible patients.

When analysed by sex, there were no significant differences in prevalence or severity of risk factors except males had an elevated mean systolic BP compared with females (difference = 12 (2 – 23) mmHg, P = 0.03).

When analysed by MHA status, there was no significant difference in prevalence or severity of risk factors, except detained patients (73%) were more likely than informal patients (41%) to be current smokers (difference = 32% (4% – 59%), P = 0.04).

The mean QRISK3 score for inpatients was significantly higher than the ‘healthy person score’ for a comparison group QRISK3 estimates with the same age, sex, and ethnicity (difference = 5.0% (3.3 – 6.7%), P < 0.001) reflecting a significantly elevated relative risk of cardiovascular disease for the inpatient group (Relative Risk = 3.8 (2.6 – 5.0)). In more meaningful terms, the mean ‘excess heart age’ (the difference between the heart age of the individual and the ‘healthy heart age’ of similar age, sex, and ethnicity) for our cohort was 11.0 (8.9 – 13.0) additional years.

When analysed by sex, males had an elevated QRISK3 relative risk compared to females (relative risk difference = 2.4 (0.1 – 4.7), P = 0.04) and this was also reflected in the increased excess heart age for males compared to females (difference = 4.6 (0.6 – 8.7) years; P = 0.02). There were no significant differences in QRISK3 outputs when analysed by MHA status.

Despite 28% (18 – 41%) of patients having a QRISK3 score of high severity (10% or greater), and 18% (10 – 30%) of patients with a QRISK 3 score of very high severity (20% of greater), only two patients (20%) of this highest severity group were prescribed lipid lowering therapy. There was no significant difference in QRISK3 severity when analysed by sex or by MHA status.

## CONCLUSIONS

### What does this study add to integrated care planning for psychiatric inpatients?

This study demonstrates that sufficient data is available to enable staff to undertake pragmatic QRISK3 estimates of cardiovascular risk for psychiatric inpatients to inform on risk reduction interventions; most of these data are readily available or obtainable during the routine admission process, and only slight modifications to admissions proforma are required to record specific information on family history of early cardiovascular disease, and more accurate delineation of smoking quantity, to enable an accurate QRISK3 assessment to be undertaken. The evaluation showed that inpatients had 3.8 times elevated QRISK3 score relative to a standardised population, and this was significantly increased in males compared to female inpatients. While nearly all patients would benefit from education and targeted support about modifiable risk factor reduction (12) (e.g. smoking, weight loss), it was found that of the 57 patients admitted who underwent assessment, 16 (28%) had a QRISK3 score greater than 10%, indicating a discussion about blood pressure and/or cholesterol lowering medication is recommended, and 10 (18%) patients had a QRISK3 score of 20% or more, indicating an urgent need to address risk factors with an offer of medication, with one patient having a QRISK3 score of 31%. This compares with the UK estimate of a high QRISK3 score (>10%) of 17.2% and very high QRISK3 score (>20%) in 8.0% of 2.6 million people in the initial QRISK3 development and validation study (14). In most cases, those patients with a QRISK3 of greater than 20% were not on lipid lowering therapy (only 2/10 (20%) patients) in the highest risk category were on any lipid-lowering therapy).

### Limitations of study

This is a small service evaluation taken from a single hospital serving a highly rural UK population, so caution is needed in extrapolation to other settings, but as a proof of concept to drive service development, it demonstrates that sufficient data is available to undertake QRISK3 assessments by rural inpatient psychiatric teams, mirroring similar efforts in other psychiatric specialty populations to improve cardiovascular health (17). Missing risk factors may reduce accuracy of QRISK3 estimates, and thus there is a need for teams to record more accurately all factors (particularly quantification of smoking, and if early family history of cardiovascular disease) to ensure accurate results, and to consider wider social factors for the individual when making assessments for intervention (18). However, we have tended to manage missing factors where mandatory for QRISK3 calculation (e.g., smoking status) by assuming lowest risk, so the results are likely to be an underestimate of cardiovascular risk. The population is an inpatient one, undergoing acute psychiatric illness often under detention, and may be less willing to offer clinical information or agree to investigations (e.g., BMI, blood pressure, and blood investigations) to allow as accurate assessment as in a primary care population, but the results shown demonstrate a high degree of routine data is collated that will allow a QRISK3 estimate to be obtained to inform on patient cardiovascular risk, and allow informed discussion with those patients who wish to engage.

### What resources are needed to enable psychiatric teams to use QRISK3 in an inpatient setting?

To implement effective integrated care to minimise cardiovascular risk in this high-risk group of patients, commissioners of mental health services will need to develop in-patient pathways and facilities with staff who have the confidence, skills, and competencies to proactively assess and offer cardiovascular risk reduction interventions to patients. This may mean having health professionals from other specialities (e.g., general practice) who are trained in QRISK3 assessment and risk factor modification reviewing each patient QRISK3 score supporting in-patient teams, or training psychiatric staff in the therapeutics of choice to reduce blood pressure and elevated lipid profiles, but we believe this is well within the capabilities of the usual psychiatric multidisciplinary team serving in-patient units, especially around behaviour modification interventions. We would suggest, however, that where possible and the patient is willing, assessment and preventive intervention is undertaken while the patient remains in hospital, rather than including this information in a discharge summary to the GP for later action, unless the patient categorically refuses. We are aware in rural areas, where a high degree of rural access deprivation for many patients with SMI exist (e.g., distance to GP surgery, lack of public transport), delaying action on cardiovascular risk modification may mean these risk factors are less likely to be addressed or opportunities missed (19). QRISK3 estimations are mainly carried out in primary care but are used less often in inpatient services; unfortunately, patients with SMI often attend and present less often to general practice to address preventable cardiovascular risk factors, and have fewer offers of intervention, less accurate coding and investigation of early cardiovascular disease and risks (20). The QRISK3 tool allows an informed discussion with patients to demonstrate how their risk can be reduced if some of the variables can be improved (this can be demonstrated in real-time with the patient), which can help support patient behaviour change. It must however be recognised that during acute admissions, many patients who are acutely unwell are not able or willing to have informed discussion about future cardiovascular risk, and these discussions may be better undertaken after the initial acute crisis has resolved during the recovery period before discharge – however, a shared care approach between primary and secondary care services (including undertaking the QRISK3 assessment annually as part of the community mental health team review of patients) to cardiovascular risk management may improve outcomes for this vulnerable patient population.

## Data Availability

All data produced in the present study are available upon reasonable request to the authors

## Funding statement

This work was supported by Health Care Research Wales via an NHS Research Time award to AW (NHS Wales RTA 21-02).

## Declarations of Interest

IB is Chief Data Scientist Advisor for AstraZeneca via the University of Liverpool Consult service, and AstraZeneca have a lipid-lowering therapeutic in their drug portfolio. There are no other authors with interests to declare.

## Data Availability

The data that support the findings of this study are available from the corresponding author (AW) upon reasonable request.

## Author Contributions

AW, LW and IB conceived the study from discussions related to research project they are contributors on around co-morbidity in patients with SMI. AW, BS, AP, MG, and JG sought study approval. AW, AP and MG collated the data. AW, LW, MB undertook the statistical analysis. All authors contributed interpretation of findings, drafting of manuscripts and final approval of materials.

## Acknowledgements

We want to thank Debbie Bufton and Caroline Johnson for their kind assistance with case records retrieval, tracking and information governance management.

## Notes

### Funding Statement

This study was supported by Health Care Research Wales via an NHS Research Time award to AW (NHS Wales RTA 21-02).

### Author Declarations

Ethics committee/IRB of Powys Teaching Health Board gave ethical approval for this work

